# Income inequality and access to advanced immunotherapy for lung cancer: the case of Durvalumab in the Netherlands

**DOI:** 10.1101/2024.04.24.24306222

**Authors:** Dimitris Katsimpokis, Mieke J. Aarts, Gijs Geleijnse, Peter Kunst, Maarten J. Bijlsma

## Abstract

With the introduction of immunotherapy with non-small lung cancer, prognosis of these patients has improved. However, socio-economic differences in access to various immunotherapy treatments have been reported. In the Netherlands, such differences are not expected due to universal insurance coverage. We investigated the existence of differential susceptibility by socioeconomic status (SES) of the effect of distance to treatment hospital on access to Durvalumab in patients with stage III non-small lung cancer who received chemoradiation, and the influence of differential mortality. We used data from the Netherlands Cancer Registry (n = 3774) from the period 2017-2021. First, we fitted Bayesian discrete failure time models and compared SES-by-distance-to-hospital interaction to a baseline model including age, distance, SES and performance score. We then fitted a time to mortality model and used both models in a g-formula to simulate a scenario where mortality levels were equalized. Our results showed that the high SES group received Durvalumab more often than the low SES group (HR = 1.23, CI_95%_ = [1.07, 1.53]), and even 4 kilometer distance increase leads to less Durvalumab (HR = 0.93, CI_95%_ = [0.86, 0.99]). Bayes factor < 3 indicated inconclusive evidence for a SES by distance interaction effect, while g-formula results showed that differential mortality does not affect SES differences. Secondary analyses showed strong evidence that SES differences in using Durvalumab were constant over the years (Bayes factor > 17). Overall, these results are significant for understanding how socio-economic inequality affects proper care and can be vital for public policy.

**What is new:**

- The higher SES group receives Durvalumab more often than the low SES group in the Netherlands.
- Increased driving distance to a Durvalumab-offering hospital lowers the probability of getting Durvalumab treatment.
- No evidence was found for SES differentials in the effect of driving distance.
- Differential mortality by SES does not affect SES differentials in Durvalumab usage.
- Bayesian hierarchical discrete failure time models capture time-to-Durvalumab data well.

## Introduction

Lung cancer is the most commonly diagnosed cancer globally, and is the leading cause of cancer-related mortality.(1) Incidence rates of lung cancer are higher among individuals with low socio-economic status (SES), and patients with low SES have poorer overall survival rates. Even after adjusting for smoking and comorbidities, this SES differential persists.(2,3) Research shows that patients belonging to lower SES groups have less access to therapy overall, not only traditional but also next-generation therapies.(4) In the Netherlands, health insurance coverage is virtually universal and does not include (economically differentiated) tiers of service for cancer therapy. This makes the Netherlands a useful case-study for causes of SES differences in access to various cancer therapies, as it eases investigation of non-institutional drivers of such differences.

Durvalumab is a recently developed immunotherapy that substantially increases progression-free survival in patients with stage III non-small cell lung cancer (NSCLC) after chemoradiotherapy.(5) The therapy was introduced in 2017 in the Netherlands for clinical studies, where it became part of the insurance package in 2019. The number of hospitals prescribing Durvalumab increased over time. Durvalumab is an expensive therapy, with its cost estimated at about 61,000 euros per patient in 2019.(6)

Two potential mechanisms of interest are travel distance to treatment facility and differential mortality. Firstly, it has been shown that increased distance to a hospital lowers probability of treatment.(7,8) However, it is unknown if this burden is greater for individuals with less economic and social capital to bridge the distance, opting instead for more traditional treatment in a less distant hospital. Secondly, it is known that as socio-economic differentials in lung cancer survival exist.(9–13) However, it is unknown if and to what degree death as a relatively stronger competing risks for individuals with lower SES explains differential access to expensive immunotherapy treatment.

The primary aim of this study is to determine whether a possible effect of distance to Durvalumab treating hospital on use of Durvalumab is moderated by SES for stage III non-small cell lung cancer (NSCLC) patients diagnosed in the period 2017-2021 in the Netherlands. To determine this, we must also establish whether SES differentials in Durvalumab usage exist, and whether a distance effect exists. The secondary aim of this study is to determine to what extent possible SES differences in use of Durvalumab is influenced by differential survival between SES groups. In addition, we evaluate whether SES modifies how use of Durvalumab unfolds across time (based on year of diagnosis) per SES group.

## Data and Methods

### Data source

Our data source is the Netherlands Cancer Registry (NCR), a population-based nationwide registry keeping records of each individual cancer patient since 1989 in the Netherlands. The registry contains demographic, tumor and primary treatment information. Information on vital status and date of death is updated annually using a computerized link with the national civil registry.

Net median household income at 6-digit postal code level in 2016 was used as proxy for SES, obtained from Statistics Netherlands. SES was clustered into 9 groups, covering 99% of the postal codes of the Dutch population. Each postal codes covers on average 17 households.

### Study population and sample

In the NCR, following clinical trials on Durvalumab effectiveness, we identified patients with clinical stage III non-small cell lung cancer (NSCLC) who received at least one cycle of chemotherapy and radiation therapy before potentially being administered Durvalumab (n=11114).(5) Patients with surgery (n=334) were excluded, as were patients with a WHO performance status score higher than 1 (n=729) or with a missing performance score (n=1208). Patients who ended up in a different (immuno)therapy regime than Durvalumab after chemoradiotherapy were also excluded (n=2040). Finally, we further excluded patients who are registered as receiving Durvalumab more than once (n = 18) as this likely indicates a registration error. The final sample size was 3774 patients.

### Outcome, censoring and competing risk

Our outcome variable is time from diagnosis until first treatment with Durvalumab, binned in 25 day units. Patients who do not receive Durvalumab by the end of follow-up are censored at the time of loss to follow-up. Death is considered a competing risk.

### Exposures

Our first main exposure is SES, categorized into 0%to 30%, 30% to 60% and 60% to 100%, representing low, medium and high SES respectively.

Our main exposure is minimum driving distance to the closest hospital providing Durvalumab (henceforth ‘driving distance’). Driving distance was calculated based on the postal code (PC4) of the patients’ residence at diagnosis and the postal code of hospitals. Availability of Durvalumab at each hospital was based on the earliest year that the therapy was recorded in the NCR. For cases of different hospital locations of the same hospital the average driving distance was taken.

We treat SES also as a potential effect modifier of the relationship between minimum-driving distance and time-to-Durvalumab treatment.

### Potential confounders and clustering

We consider age at diagnosis, WHO performance status score and year of diagnosis as potential confounding variables. Age was measured in years. Performance status followed the WHO scale, where ‘0’ indicates asymptomatic patients and ‘1’ indicates symptomatic patients but fully ambulant. Year of diagnosis was based on semesters (defined from January to June, and from July to December) and treated as a continuous variable. Furthermore, we consider observations to be clustered within the hospital which the patients first visited (commonly hospital of diagnosis and referral) with symptoms related to lung cancer.

### Statistical analysis

Our estimands of interest are the restricted mean survival times (RMSTs) if all individuals had low, medium, and high SES, and the differences between these quantities, as well as the marginal hazard ratios between these groups. These quantities were also calculated for percentiles of distance from hospital (10%, 30%, 50%, 70% and 90%) as compared to a distance of zero kilometers from the hospital.

Furthermore, to gain insight into the influence of death as an outcome that would prevent receiving Durvalumab treatment, also known as a competing risk, we also approximated the SES estimand under a scenario where death distributions were equalized between SES groups. We set all groups to have the mortality levels, conditional on their other covariates, of the low SES group. To do so, we first assume the extended DAG shown in Figure 1. Secondly, following this DAG, we fit a Bayesian hierarchical discrete failure time model (DFT) with time to Durvalumab as the outcome variable, and the minimum driving distance, SES, and potential confounders as covariates. Inclusion of an interaction term between SES and driving distance, and between diagnostic year and SES, were determined using Bayes factor with a factor of 3 as the cutoff for inclusion.(14,15) Patients are assumed nested within hospitals of first contact. This procedure was also followed for a second DFT with time to death as the outcome variable. These results from these models were used in a g-formula with Monte Carlo integration to approximate the aforementioned quantities of interest.(16) The modelling procedure is described in more detail in appendix A. In the distance estimand, we used the observed death distribution, i.e. without equalizing death.

**Figure 1.**
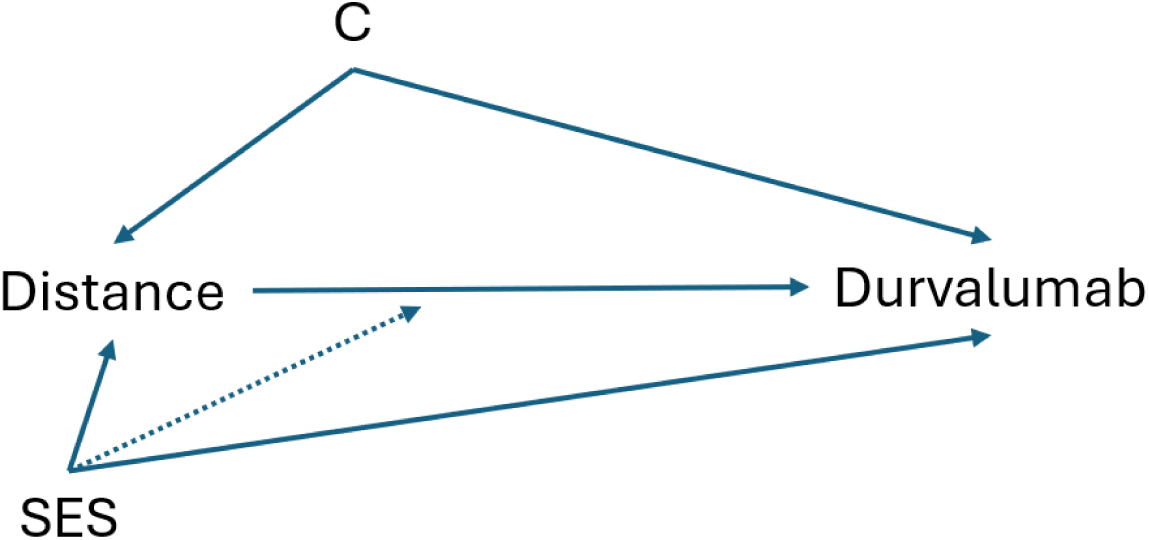
Extended Directed Acyclic Graph (DAG) of the assumed causal relationships between variables in our study. ‘SES’ stands for ‘socio-economic status’, while ‘C’ stands for the potential confounders age at diagnosis, WHO performance status score, and year of diagnosis, which are not plotted separately for brevity. Interaction effects are shown by dotted lines as they violate basic DAG principles. The interaction between year of diagnosis and SES is further not shown for simplicity.

## Results

Table 1 shows the descriptive statistics of the variables included in the statistical models. Of interest is that 37% of included patients received Durvalumab, and that 10% of patients died before they could be prescribed Durvalumab within the follow-up window of 250 days since diagnosis. Within this window, the average time-to-receive Durvalumab was 160 days and the average time to death for our cohort was 168 days. Most included patients, 54% lived in a neighborhood with a medium SES, as compared to low (27%) and high (17%).

**Table 1.** Descriptive cohort data (n = 3774). ‘SD’ indicates ‘standard deviation’. The table is limited to observations up to 250 days, which is end of follow-up in this study.

| Variable | Counts/percentages/averages |
| --- | --- |
| Received Durvalumab | Yes = 1388 (37%); No = 2386 (63%) |
| Vital status | Dead = 398 (10%); Alive = 3367 (90%) |
| Time To Durvalumab (mean days) | 160.4 (SD = 56.9) |
| Time to Death (mean days) | 168.9 (SD = 56.2) |
| Age (mean years) | 66.1 (SD = 8.9) |
| WHO Performance status | 0 = 1968 (52%); 1 = 1896 (47%) |
| SES | Low = 1028 (27%); Medium = 2099 (56%); High = 647 (17%) |
| Driving distance (km) | 21.2 (SD = 22.8) |
| Year of diagnosis (a = Jan-June, b = July-Dec) | 2017a = 325 (8%); 2017b = 351 (9%) |
|  | 2018a = 422 (11%); 2018b = 399 (10%) |
|  | 2019a = 425 (11%); 2019b = 385 (10%) |
|  | 2020a = 337 (8%); 2020b = 381 (10%) |
|  | 2021a = 354 (9%); 2021b = 391 (10%) |

Table 2 presents the results of the model comparison to determine if interactions between SES and driving distance, and between SES and year of diagnosis, should be included. We found neither evidence for nor against the inclusion of the interaction term between SES and driving distance; the baseline model was

**Table 2.** Model comparison (time to Durvalumab) of two nested models against the baseline through Bayes Factors. Parameter difference indicates the number of additional free parameters of each nested model as compared to the baseline model. Bayes factors indicate likelihood ratio of the baseline model over the corresponding nested one. The Equations of the models are given in appendix A. Time to death model comparison is found in appendix B.

| Model | Parameter difference | Bayes factor |
| --- | --- | --- |
| Baseline | — | — |
| Baseline + SES*driving distance | 2 | 2.4 |
| Baseline + SES*year of diagnosis | 2 | 17.1 |

2.4 times more likely than the model with interaction (3 was the decision threshold). Furthermore, we found strong evidence against the inclusion of the interaction term between SES and year of diagnosis; the baseline model was 17.1 times more likely. The comparison between the natural course simulation from the baseline model and the empirical Kaplan Meier curve showed some underestimation of survival in days 50 to 100, but no major divergences (appendix C).

The coefficient of year of diagnosis indicated a positive association (OR = 2.22, CI_95%_ = [2.08, 2.38]). Additionally, our covariates of performance status and age both showed a negative association with respect to receiving Durvalumab (OR = 0.71, CI_95%_ = [0.62, 0.80], and OR = 0.80, CI_95%_ = [0.76, 0.85] respectively).

To formally test the effect of SES group (corresponding to the average treatment effect of SES), we calculated Hazard Ratios (HR) and Restricted Mean Survival Time (RMST) based on the time-to-Durvalumab probability curves of the counterfactual scenario B simulation (see Table 3). Overall, both HRs and RMSTs suggested that the high SES group receives Durvalumab earlier than the low SES group. HRs and RMSTs based on scenario A derived very similar values to those shown in Table 3 from scenario B.

**Table 3.**
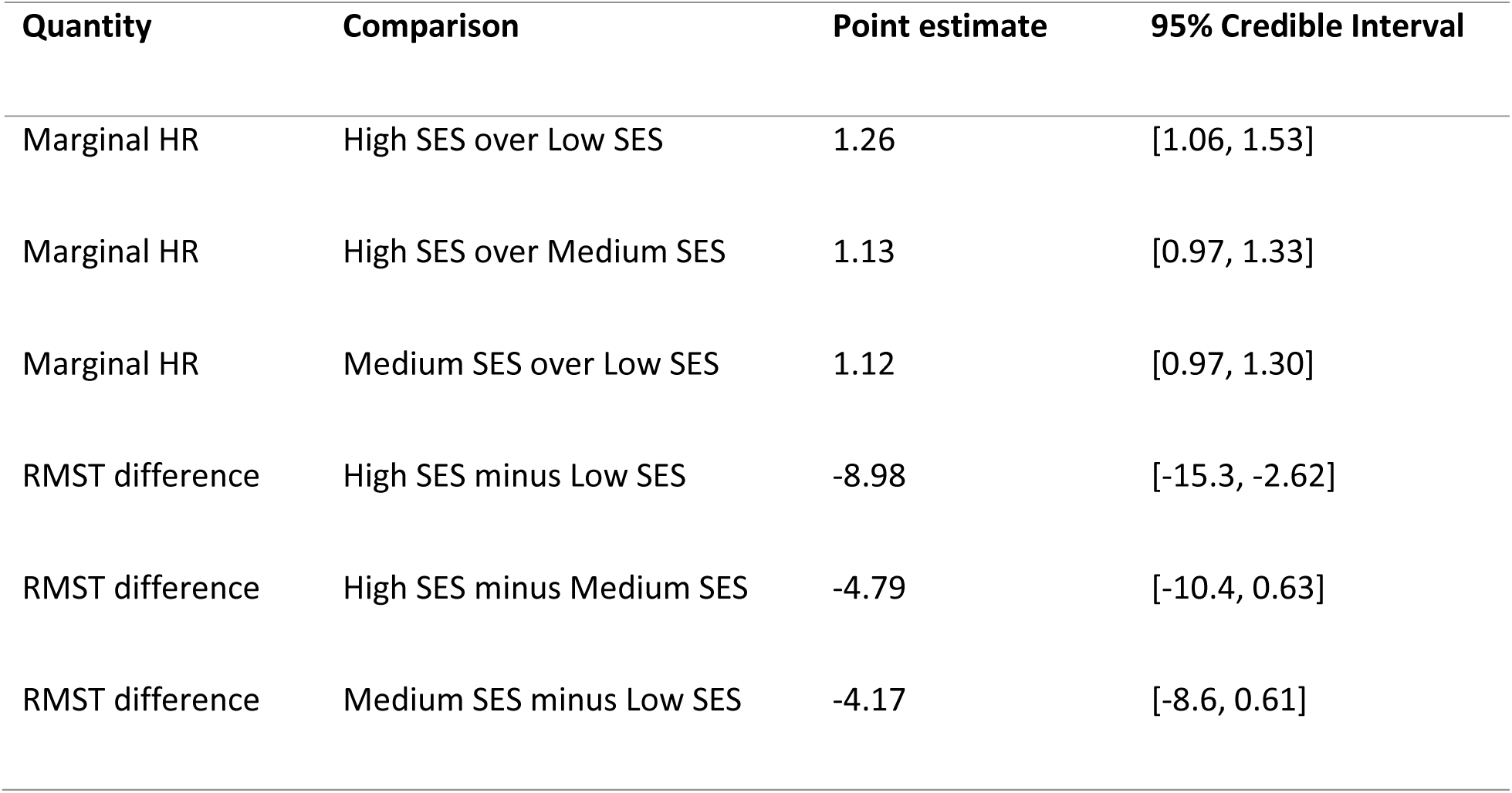
Average SES effect based on scenario B (counterfactual, equalized death). The credible interval is calculated based on 2.5% and 97.5% quantile. ‘HR’ stands for Hazard Ratio, and ‘RMST’ for restricted mean survival time.

| Quantity | Comparison | Point estimate | 95% Credible Interval |
| --- | --- | --- | --- |
| Marginal HR | High SES over Low SES | 1.26 | [1.06, 1.53] |
| Marginal HR | High SES over Medium SES | 1.13 | [0.97, 1.33] |
| Marginal HR | Medium SES over Low SES | 1.12 | [0.97, 1.30] |
| RMST difference | High SES minus Low SES | -8.98 | [-15.3, -2.62] |
| RMST difference | High SES minus Medium SES | -4.79 | [-10.4, 0.63] |
| RMST difference | Medium SES minus Low SES | -4.17 | [-8.6, 0.61] |

Regarding the effect of driving distance (see Table 4), both HRs and RMSTs indicated that even a small increase of distance (about 4 kilometers corresponding to the bottom 10% of the distance distribution) compared to zero distance leads patients to receive Durvalumab less and later, with estimates changing non-linearly with more distance increments (see Appendix D for more details).

**Table 4.** Average driving distance effect based on scenario A (counterfactual, observed [non-equalized] death). Comparisons are based on five quantiles of the observed distance distribution vs. zero kilometer distance from the nearest Durvalumab-providing hospital. The corresponding kilometer values are given next to each quantile level. The credible interval is calculated based on 2.5% and 97.5% quantile of the posterior predictive distribution. ‘HR’ stands for Hazard Ratio, and ‘RMST’ for restricted mean survival time, ‘KM’ for ‘kilometer’ and ‘q’ for ‘quantile’.

| Quantity | Comparison | Point estimate | 95% Credible Interval |
| --- | --- | --- | --- |
| Marginal HR | 3.91 KM (10% q) over 0 KM | 0.93 | [0.86, 0.99] |
| Marginal HR | 8.77 KM (30% q) over 0 KM | 0.86 | [0.77, 0.91] |
| Marginal HR | 14.8 KM (50% q) over 0 KM | 0.77 | [0.67, 0.85] |
| Marginal HR | 23.3 KM (70% q) over 0 KM | 0.68 | [0.55, 0.76] |
| Marginal HR | 43.7 KM (90% q) over 0 KM | 0.46 | [0.33, 0.59] |
| RMST difference | 3.91 KM (10% q) minus 0 KM | 2.99 | [1.84, 4.17] |
| RMST difference | 8.77 KM (30% q) minus 0 KM | 6.58 | [4.45, 8.97] |
| RMST difference | 14.8 KM (50% q) minus 0 KM | 10.91 | [7.52, 14.64] |
| RMST difference | 23.3 KM (70% q) minus 0 KM | 16.57 | [11.60, 22.09] |
| RMST difference | 43.7 KM (90% q) minus 0 KM | 28.55 | [20.48, 37.01] |

Additionally, early mortality equalization did not reveal any SES differentials and, therefore results are shown in Appendix E.

## Discussion

The primary aim of this study is to determine whether a possible effect of distance to Durvalumab treating hospital on use of Durvalumab is moderated by SES for stage III non-small cell lung cancer (NSCLC) patients diagnosed in the period 2017-2021 in the Netherlands. Our findings neither support nor negate effect modification of SES on driving distance. Before establishing this finding, we first had to establish main effects of SES and driving distance. Firstly, we found a strong negative effect of driving distance: 7% lower hazard to receive Durvalumab with a distance increase from zero kilometers to just the bottom 10% of observed distance (i.e., about 4 kilometers distance), with hazards being 16%, 23%, 32% and 54% lower for Durvalumab corresponding to scenarios where distance is set to the bottom 30%, 50%, 70% and 90% of observed distance as compared to zero kilometers. Secondly, in the scenarios where all patients were set to high, medium and low SES we found that in the high SES scenario had a 26% higher hazard to receive Durvalumab than in the low SES scenario (CI_95%_ 1.07 to 1.53).). As a secondary analysis, we determined whether the effect of SES on Durvalumab can be (partly) explained by the differential early mortality of lower income earners, which could, in turn, prevent use of Durvalumab. Our findings suggest that the effect of SES on using Durvalumab after equalizing the death distribution of the different SES groups was very similar to the effect given the observed death distributions. Finally, although we found an overall increase of the usage of Durvalumab from 2017 to 2021 by about 60% (OR = 2.22, CI_95%_ = [2.08, 2.38]), we found strong evidence against effect modification of SES on year of diagnosis, indicating no evidence for closing or increasing the gap between high SES and low SES over the years. Overall, these findings suggest that the direct effect of SES on using Durvalumab cannot be explained away by either the differential effect of driving distance to the closest center offering Durvalumab, or by differential survival.

### Evaluation of data and methods

Our study has a number of strengths. Firstly, internal validity is strong as the number of relevant confounding variables is limited; conditional on SES, mobility decisions tend to be taken with distance to work or family in mind and not with distance to a hospital (specifically hospital treating with Durvalumab) in mind.(17) Few variables exist that influence distance to such a hospital while also affecting treatment decisions regarding Durvalumab. The number of hospitals that provide Durvalumab treatment was greatly expanded in our data set, starting at 10 hospitals in 2017 to 72 in 2021, strengthening the exogeneity of the distance variable. In addition, we covered all confounding variables following the medical guidelines in the Netherlands on which patients may receive Durvalumab and who may not.

Secondly, we consider the functional form of our DFT model as an additional strength. We provided a flexible functional (natural spline) form of the time predictor to account for the non-linear effect of time.(18) This spline effectively models the equivalent of what would be the baseline hazard curve in other common time-to-event models. We opted for linear relationships for the rest of the covariates and the distance exposure for simplicity as well as after having visually examined the resulted (quasi-linear) form of such relationships when applying a spline transformation to them.

Finally, death is a competing risk for time-to-Durvalumab. By censoring patients at the time of death, it is implicitly assumed that they would have the same hazards of receiving Durvalumab as another individual with the same covariate values still alive at that time. However, this represents an untestable assumption. For this reason, in our equalized death scenario (scenario B), we gave all individuals the survival probabilities of individuals with low SES, conditional on their other covariate values; this represents a scenario in which we made medium and high SES individuals die sooner, rather than later, thereby avoiding the aforementioned untestable assumption. As the authors are unaware of similar work, we note that this may be the first study to perform such a counterfactual experiment to determine the possible influence of competing risks on our primary research aim, hence some caution is warranted in interpreting these results.

An additional possible limitation of our work consists of the exclusion of a sizeable (n=2040) number of patients who were given a different (immuno)therapy regime than Durvalumab after receiving chemoradiotherapy. Systematic variation of ending up in either of these therapy regimes could be a potential bias for our results: for example, if patients belonging to the low SES group were systematically given other (immuno)therapy regimes than Durvalumab (and vice-versa). Given that the choice between other (immuno)therapies and Durvalumab is mutually exclusive, future research could go a step further by equalizing the reception of immunotherapy distribution (thereby, treating the reception of other (immuno)therapies as a competing risk). However, at this time there is no evidence that the low SES group has higher usage (than other SES groups) of other types of immunotherapy.

Finally, a limitation concerns the follow-up period studied; patients are selected if they completed at least one round of chemotherapy and radiation therapy. However, our follow-up time starts at diagnosis. This effectively means that patients are functionally immortal until they finish their first round of chemotherapy and radiation therapy(19). Differential delays in reaching these inclusion conditions could affect timing of first Durvalumab treatment as well. In this paper, we effectively study the accumulation of SES differences in both such a potential delay and the one to Durvalumab. Future research should determine the degree to which SES differences in time-to-Durvalumab are explained by initial differences in time-to-first chemotherapy and radiotherapy between the SES groups.

### Connection to previous studies

Our study showed patients of higher SES tend to have more use of immunotherapy for NSCLC. This is in line with previous studies showing similar results for treatments in other cancer types.(20,21) Research has shown the general trend of elevated end-of-life costs for higher SES groups.(22)

In addition, our finding of a negative association between administration of Durvalumab and driving distance to the closest Durvalumab-offering hospital can be seen in the broader context of regional variations of cancer treatment in the Netherlands. For example, regional variations have been observed in the use of radiotherapy for prostate cancer(23) and rectal cancer(24). Specifically for NSCLC, recent evidence suggests that regional variations in the Netherlands are associated with differential use of radiotherapy and type of chemoradiotherapy (i.e., sequential vs. concurrent).(7) Finally, our data remain inconclusive regarding modification of the effect of driving distance by SES, as shown by Bayes Factors and parameter estimates.

### Further possible pathways

There are many possible pathways through which SES affects use of Durvalumab. For example, individuals with a higher SES tend to have more developed social capital, which can make them more informed or aware of new cancer therapies. The relationship between SES and health literacy is well documented in the literature.(25–27) This, in turn, may cause individuals with a higher SES to be more demanding of the latest and advanced therapies as compared to individuals with a low SES. One way to indirectly test this pathway would be to assess if individuals with a high SES switch hospitals of therapy between diagnosis and administration of Durvalumab, and therefore, bypassing the initial plan of treatment provided by the medical group at the time of diagnosis.

Moreover, other studies have stressed the link between race, ethnicity and SES as a factor for receiving appropriate care and treatment, not only in lung cancer(12), but also more generally.(28) For example, in the Netherlands non-Western ethnic groups have been shown to receive adjuvant chemotherapy for colon cancer less, which cannot be explained away by SES.(29) Therefore, a potentially mutual influence between SES and ethnicity and their combined effect on Durvalumab (or other immunotherapies) could be studied in future research.

Finally, in this study, following clinical trial inclusion criteria, we focused on a patient cohort with good performance status, and thereby excluding patients with likely multimorbidity. Future research could pay more attention to the interplay between multimorbidity and receiving immunotherapy for lung cancer, given that evidence suggests that multimorbidity and ethic differences are related and not fully explained away by SES.(30) Similar findings have been reported worldwide for a variety of clinical factors.(31,32)

## Conclusion

Our work provides evidence that patients with high SES are more likely to receive the immunotherapy Durvalumab in the Netherlands for the treatment of stage III NSCLC, after chemoradiation. We showed that this effect cannot be explained by differential early mortality of patients by socio-economic status (SES). Our data remain inconclusive regarding the differential effect of driving distance by SES.

## Data Availability

Data are not directly available, but need to be requested from Integraal Kankercentrum Nederland (IKNL) through a formal application procedure (www.iknl.nl)

## Statements and Declarations

### Ethics approval

According to the Dutch Central Committee on Research involving Human Subjects, no ethical approval is needed for this study, as it is a retrospective study, which uses data from the NCR.

